# A Framework for Combining Morbidity and Mortality to Identify Determinants of Child Health

**DOI:** 10.1101/2025.09.19.25336127

**Authors:** Anuradha R Chetiya, Vishal Deo

## Abstract

**Objective:** The objective of this study is to combine morbidity in terms of various levels of growth faltering or malnutrition among surviving children with mortality to better understand the determinants of infant and child health.

**Method:** This study uses data from the latest National Family Health Survey (NFHS)-5 of India conducted during 2019-21 to provide a framework that integrates both mortality and morbidity as a measure of infant and child health to investigate their association with maternal and other socio-economic determinants of health. The outcome variable representing health status of a child is defined as an ordinal variable with four progressively worsening levels– no malnutrition, moderate malnutrition, severe malnutrition, and not alive. The levels of morbidity in terms of growth faltering or malnutrition are defined in line with the World Health Organization (WHO) guidelines. Multivariable ordinal logistic regression has been used to assess the association of risk factors with the outcome variable.

**Results:** Among the examined determinants, maternal education and household wealth index demonstrated the strongest associations with child health outcomes. Notably, the risk of adverse health outcomes did not significantly differ between male and female children, indicating gender parity in child health—a positive development for the country’s public health landscape.

**Conclusion:** Combining child mortality and morbidity allows better indication of child health status with added sensitivity through progressive health status levels, ranging from best scenario (no malnutrition) to worst scenario (death). Consequently, it enables a more comprehensive assessment of maternal and socioeconomic determinants of child health. In addition, using such combined outcome ensures complete utilization of information available in a survey dataset, irrespective of the mortality status of the child.

## 1. Introduction

Poor health in early childhood, including malnutrition or infections, can lead to chronic diseases in adulthood affecting the quality of life of an individual. Research studies have linked health of children in their early developmental age to non-communicable risk factors at the later stage of their life.^1-6^Early childhood represents a window of opportunity for interventions like improving nutrition, vaccination, and healthcare access to ensure a healthier life. Investing in early childhood health lowers the economic burden on a country’s healthcare systems by preventing chronic illnesses that are often expensive to manage later in adulthood.

Research studies on infant and child health have focused on identifying determinants of the standard mortality rates like IMR (infant mortality rate), under five mortality rate and similar measures. ^2,7-13^While mortality is the number of deaths and is absolute, morbidity is a measure of those affected by the health condition and is not absolute as there are chances that some may survive while others may not, or some may survive but with lower quality of health. So, it becomes pertinent that child health status be examined not just in the terms of mortality, but also by incorporating child growth parameters as outcomes of interest, which can together present a more comprehensive picture. The Mosley Framework^14^is a seminal analytical model designed to study child health, mortality, and morbidity. Developed by Henry Mosley and Lincoln Chen in 1984, it integrates social, economic, and biological factors to identify determinants of child health outcomes. The analytical framework proposed by Mosley and Chen (1984) for child survival provides a basis for understanding the many factors that influence bringing up of a healthy child in a developing economy. While social scientists tend to focus on mortality, most medical scientists typically tend to focus on health status of survivors of a disease. Mosley and Chen (1984) argued that focusing merely on mortality ‘handicaps research as death is a rare event’. They suggested integrating both approaches by combining counts of the dead with observations on the living to obtain a unified scale or index for determining the health status of a population. Based on the premise that, ‘in an optimal setting over 97 percent of newborns can be expected to survive through the first five years of life’, their framework provides a structured approach to understanding how interventions, such as improved healthcare, nutrition, or sanitation, can reduce child mortality and morbidity.

The present study draws motivation from the idea given by the Mosley-Chen framework. It combines morbidity in terms of various levels of growth faltering or malnutrition among surviving children, with mortality to better understand the determinants of infant and child health. This study uses data from the latest National Family Health Survey (NFHS) -5^15^of India conducted during 2019-21, to provide a framework that incorporates both mortality and morbidity as a measure of infant and child health to investigate their relation to maternal and other socioeconomic determinants of health. While using demographic health survey data, like NFHS, taking mortality status of children as the outcome measure leads to non-utilization of information on children who may be alive but not healthy. In addition, the presence of censoring with respect to mortality outcome adds to the limitation of such analysis. The NFHS 5 provides data of children up to five years of age. So, for example, if the event of interest is under-five mortality, information on a child who is alive and less than five years of age at the time of the survey is censored as the event may occur in the future. Our framework overcomes both these limitations by incorporating health status of all children irrespective of their mortality status.

## 2. Methodology

### 2.1. Data and Sample Design

This analysis utilizes data from the fifth round of the NFHS conducted during 2019-21 in India. The NFHS is a nationally representative cross-sectional survey that collects data on a large number of demographic, socioeconomic, maternal and child health parameters. The previous four rounds of the NFHS were conducted in 1992-1993 (NFHS-1)[16], 1998-1999 (NFHS-2)[17], 2005-2006 (NFHS-3) and 2015-2016 ( NFHS-4). The NFHS surveys were carried out under the guidance of the Ministry of Health and Family Welfare (MoHFW), Government of India with the International Institute for Population Sciences (IIPS), Mumbai, as the nodal agency. The overall sample size for NFHS-5 was based on several factors, the most important being the requirement to generate indicators at the district and/or state and union territory (UT) levels. NFHS-5 was conducted in 707 districts, 28 states, and 8 union territories. In this round of NFHS 5, a two-stage stratified sampling approach has been used. Each district was divided into urban and rural strata. From each rural sampling stratum, villages were selected as Primary Sampling Units (PSUs). For the urban strata, Census Enumeration Blocks (CEBs) were selected as PSUs. A total of 664,972 households were selected for the sample, out of which 653,144 were found to be occupied. Interviews were successfully conducted in 636,699 of these households, yielding a response rate of 98 percent. Within the interviewed households, 747,176 women aged 15–49 were identified as eligible for individual interviews, and interviews were successfully completed with 724,115 women, resulting in a response rate of 97 percent.^15^Data from the children recode file was used in this analysis.

### 2.2. Dependent Variable

To incorporate both mortality and morbidity in the analysis, four levels of the dependent variable were defined. Three levels of morbidity in terms of growth faltering or malnutrition were defined in line with the World Health Organization (WHO) guidelines.The WHO classification for malnutrition considers standard deviation or Z scores of weight for length/ height for classifying children in moderate (between -2 to -3 SD) and severe(<-3SD) categories.^16^ For our analysis the four levels of the dependent variable that we have considered are, 1: No Malnutrition(greater than -2SD), 2: Moderate Malnutrition(between -2 to -3SD), 3: Severe Malnutrition(less than - 3SD), and4: Not Alive.That is, the dependent variable level 1 represents the best possible outcome and level 4 represents the worst possible outcome.

### 2.3. Independent Variables

Socioeconomic and demographic factors identified in various studies include nutritional status of mother, age of the mother, gaps between two deliveries, mother’s education level, access to healthcare services that ensure safe delivery along with antenatal and postnatal care.^2,8-12,17^Since maternal health is an immediate and principal factor in determining child mortality, we classified one category of independent variables under the umbrella of maternal factors and the second category of independent variables under environmental factors. This classification follows the broader framework of socioeconomic determinants that influence risk of mortality and morbidity in a developing countrygiven byMosley and Chen (1984). Data on all variables included as predictor variables in the study are available in the child recode file.

#### 2.3.1. Maternal Variables

The following maternal variables were considered for analysis.

##### Age of the mother at the time of birth

For our analysis we have classified current age of mother under three categories –15-24 years, 25-34 years and 35 – 49 years using data on mother’s age from the child recode file.

##### Birth order

Birth order of the child considered in the study was classified into three groups viz. first, second or third, fourth or more.

##### Full AnteNatalCare (ANC) Utilization

A mother is considered to have received full ANC (ante natal care) if she received minimum required TT dosages, consumed prophylactic IFA supplement for at least 100 days, and completed four or more ANC visits is considered to have received full ANC.

##### Place of delivery

The NFHS compiles data on the place of delivery across 17 categories, encompassing delivery at the respondent’s home, another home, a parent’s home, hospitals, public health facilities, primary health centers, dispensaries, clinics, as well as both private and public healthcare facilities. All deliveries other than respondent’s home, other home and parent’s home have been considered as institutional deliveries. And all home deliveries have been taken as non-institutional deliveries.

##### Educational level of the mother

In India, the school education system comprises 10+2 years of schooling, including ten years of primary and secondary education followed by two years of higher secondary education.For this variable, four categories of education level were defined – mothers with no education, mothers who have completed primary education, secondary education and higher education respectively.

##### Birth weight

A child under 2500 grams has been considered as underweight and all children above this weight has been considered as having normal birth weight. This is according to the WHO definition of ‘low birth weight’.

#### 2.3.2. Environmental Variables

The following variables have been included in the category of environmental variables for the analysis.

##### Wealth index

The NFHS-5 survey assessed the economic status of households using a measure called the wealth index. This index assigned scores to households based on the quantity and type of consumer goods they owned, such as a television, bicycle, car, or motorcycle, as well as housing characteristics like the source of drinking water, toilet facilities, and flooring type. Households were then categorized into five wealth quintiles, each representing 20% of the population. All five quintiles (lowest, second, middle, fourth, and highest) have been included in the analysis.

##### Place of residence

The types of place of residence have been taken as rural and urban households. A total 14816 urban and 42954 rural households have been included in the analysis.

###### Cooking fuel

NFHS data is available for more than 10 categories of cooking fuel used. For this analysis we have considered electricity, liquefied petroleum gas, natural gas and biogas as safe fuels and the remaining fuel sources which includes kerosene, charcoal, wood, animal dung and agricultural crop among others as unsafe fuels.

##### Sex of the child

Whether the child is male or female are the two categorizations for this variable. The environment significantly influences gender development in children specially in developing economies. Social, cultural, and economic factors often shape gender roles and expectations from an early age. Issues like poverty, urban crowding, and lack of infrastructure can exacerbate gender inequalities particularly affecting girls thereby impacting their growth and mental health.

### 2.4. Statistical Analysis

The total number of children upto five years of age in the child recode file were 87264. Out of these, 57,770 children have been included in the study after accounting for missing observations across all variables. Data has been analyzed using STATAsoftware version 18. Bivariate association between each factor and the outcome variable was assessed using Chi-square test for independence of attributes. A summary of the maternal and environmental factors and their bivariate association with the dependent variable is provided in Table 1.

**Table 1.** Bivariate associations of maternal and environmental factors with the dependent variable.

| Summary of Maternal Factors |  |  |  |  |  |  |
| --- | --- | --- | --- | --- | --- | --- |
|  | No Malnutrition | Moderate malnutrition | Severe malnutrition | Not alive | Total | Test |
| Total | 34637 | 17674 | 2340 | 3119 | 57,770 |  |
| Mother's current age |  |  |  |  |  |  |
| 15-24 | 9,125 (26.3%) | 4,617 (26.1%) | 374 (16.0%) | 1,017 (32.6%) | 15,133 (26.2%) | <0.001 |
| 25-34 | 21,839 (63.1%) | 10,863 (61.5%) | 1,498 (64.0%) | 1,674 (53.7%) | 35,874 (62.1%) |  |
| 35-49 | 3,673 (10.6%) | 2,194 (12.4%) | 468 (20.0%) | 428 (13.7%) | 6,763 (11.7%) |  |
| Birth Order |  |  |  |  |  |  |
| First | 12,687 (36.6%) | 5,175 (29.3%) | 563 (24.1%) | 975 (31.3%) | 19,400 (33.6%) | <0.001 |
| Second or third | 19,103 (55.2%) | 9,143 (51.7%) | 1,259 (53.8%) | 1,462 (46.9%) | 30,967 (53.6%) |  |
| Fourth or more | 2,847 (8.2%) | 3,356 (19.0%) | 518 (22.1%) | 682 (21.9%) | 7,403 (12.8%) |  |
| Full ANC status |  |  |  |  |  |  |
| No | 21,680 (62.6%) | 12,706 (71.9%) | 1,638 (70.0%) | 2,357 (75.6%) | 38,381 (66.4%) | <0.001 |
| Yes | 12,957 (37.4%) | 4,968 (28.1%) | 702 (30.0%) | 762 (24.4%) | 19,389 (33.6%) |  |
| Highest educational level of mother |  |  |  |  |  |  |
| No education | 3,461 (10.0%) | 4,944 (28.0%) | 775 (33.1%) | 892 (28.6%) | 10,072 (17.4%) | <0.001 |
| Primary education | 2,907 (8.4%) | 2,650 (15.0%) | 338 (14.4%) | 478 (15.3%) | 6,373 (11.0%) |  |
| Secondary education | 19,729<br>(57.0%) | 8,629 (48.8%) | 1,063 (45.4%) | 1,478<br>(47.4%) | 30,899<br>(53.5%) |  |
| Higher education | 8,540 (24.7%) | 1,451 (8.2%) | 164 (7.0%) | 271<br>(8.7%) | 10,426<br>(18.0%) |  |
| Delivery by caesarean section |  |  |  |  |  |  |
| No | 21,853<br>(63.1%) | 15,195 (86.0%) | 1,988 (85.0%) | 2,478<br>(79.4%) | 41,514<br>(71.9%) | <0.001 |
| Yes | 12,784<br>(36.9%) | 2,479 (14.0%) | 352 (15.0%) | 641<br>(20.6%) | 16,256<br>(28.1%) |  |
| Birth Weight |  |  |  |  |  |  |
| Low birth weight | 4,779 (13.8%) | 3,713 (21.0%) | 545 (23.3%) | 1,084<br>(34.8%) | 10,121<br>(17.5%) | <0.001 |
| Normal birth weight | 29,858<br>(86.2%) | 13,961 (79.0%) | 1,795 (76.7%) | 2,035<br>(65.2%) | 47,649<br>(82.5%) |  |
| **Summary of Environmental Factors** |  |  |  |  |  |  |
|  | No<br>Malnutrition | Moderate<br>malnutrition | Severe<br>malnutrition | Not alive | Total | Test |
| Type of place of residence |  |  |  |  |  |  |
| Urban | 11,534<br>(33.3%) | 2,428 (13.7%) | 318 (13.6%) | 536<br>(17.2%) | 14,816<br>(25.6%) | <0.001 |
| Rural | 23,103<br>(66.7%) | 15,246 (86.3%) | 2,022 (86.4%) | 2,583<br>(82.8%) | 42,954<br>(74.4%) |  |
| Cooking Fuel |  |  |  |  |  |  |
| Safe | 23,130<br>(66.8%) | 6,061 (34.3%) | 825 (35.3%) | 1,197<br>(38.4%) | 31,213<br>(54.0%) | <0.001 |
| Unsafe | 11,507<br>(33.2%) | 11,613 (65.7%) | 1,515 (64.7%) | 1,922<br>(61.6%) | 26,557<br>(46.0%) |  |
| Place of delivery |  |  |  |  |  |  |
| Non- institutional | 1,343 (3.9%) | 1,793 (10.1%) | 231 (9.9%) | 297 (9.5%) | 3,664<br>(6.3%) | <0.001 |
| Institutional | 33,294<br>(96.1%) | 15,881 (89.9%) | 2,109 (90.1%) | 2,822<br>(90.5%) | 54,106<br>(93.7%) |  |
| Wealth index combined |  |  |  |  |  |  |
| Poorest | 3,139 (9.1%) | 6,610 (37.4%) | 939 (40.1%) | 991<br>(31.8%) | 11,679<br>(20.2%) | <0.001 |
| Poorer | 5,572 (16.1%) | 4,646 (26.3%) | 592 (25.3%) | 839<br>(26.9%) | 11,649<br>(20.2%) |  |
| Middle | 7,355 (21.2%) | 3,178 (18.0%) | 399 (17.1%) | 558<br>(17.9%) | 11,490<br>(19.9%) |  |
| Richer | 8,878 (25.6%) | 2,148 (12.2%) | 274 (11.7%) | 474<br>(15.2%) | 11,774<br>(20.4%) |  |
| Richest | 9,693 (28.0%) | 1,092 (6.2%) | 136 (5.8%) | 257 (8.2%) | 11,178<br>(19.3%) |  |
| The child's gender |  |  |  |  |  |  |
| Male | 18,647<br>(53.8%) | 9,609 (54.4%) | 1,267 (54.1%) | 1,773<br>(56.8%) | 31,296<br>(54.2%) | 0.013 |
| Female | 15,990<br>(46.2%) | 8,065 (45.6%) | 1,073 (45.9%) | 1,346<br>(43.2%) | 26,474<br>(45.8%) |  |

Since the proposed dependent variable has ordinal scale, multivariable ordinal logistic regression was fitted to find adjusted odds ratios associated with all factor variables included in the study. The model was fitted while accounting for survey weights. Results from the multivariable ordinal logistic regression analysis are presented in Table 2.

**Table 2.** Results of the multivariable ordinal logistic regression.

| Factors/ Covariates | Odds ratio | Std. Error | t | P>\|t\| | [95% conf. interval] |  |
| --- | --- | --- | --- | --- | --- | --- |
| Maternal Factors |  |  |  |  |  |  |
| Mother's current age [Baseline: |  |  |  |  |  |  |
| 25-34 | 1.131192 | .0379315 | 3.68 | 0.000 | 1.059234 | 1.208038 |
| 35-49 | 1.269216 | .0719557 | 4.21 | 0.000 | 1.135732 | 1.418388 |
| Birth Order |  |  |  |  |  |  |
| Second or third | .8555318 | .0262121 | -5.09 | 0.000 | .8056667 | .9084831 |
| Fourth or more | 1.020519 | .0463434 | 0.45 | 0.655 | .9336084 | 1.11552 |
| Full ANC status |  |  |  |  |  |  |
| Yes | .8656938 | .0241086 | -5.18 | 0.000 | .8197061 | .9142616 |
| Highest educational level of mother |  |  |  |  |  | .8354027 |
| Primary education | .7725917 | .0308094 | -6.47 | 0.000 | .7145032 | .8354027 |
| Secondary education | .6411433 | .0204974 | -13.90 | 0.000 | .6022 | .682605 |
| Higher education | .4784285 | .0290208 | -12.15 | 0.000 | .4247973 | .5388306 |
| Birth Weight |  |  |  |  |  |  |
| Normal birth weight | .6032518 | .0193923 | -15.72 | 0.000 | .5664145 | .6424849 |
| Environmental Factors |  |  |  |  |  |  |
| Type of place of residence |  |  |  |  |  |  |
| Rural | 1.273999 | .0577189 | 5.35 | 0.000 | 1.165744 | 1.392307 |
| Cooking Fuel |  |  |  |  |  |  |
| Unsafe | 1.172987 | .0368062 | 5.08 | 0.000 | 1.103018 | 1.247394 |
| Place of delivery |  |  |  |  |  |  |
| Institutional | .8318941 | .0394196 | -3.88 | 0.000 | .7581089 | .9128607 |
| Wealth index combined |  |  |  |  |  |  |
| Poorer | .645953 | .0213602 | -13.22 | 0.000 | .6054137 | .6892069 |
| Middle | .408166 | .0161629 | -22.63 | 0.000 | .3776839 | .4411082 |
| Richer | .2829143 | .0133996 | -26.66 | 0.000 | .2578325 | .3104362 |
| Richest | .1542364 | .0109754 | -26.27 | 0.000 | .1341568 | .1773214 |
| The child's gender |  |  |  |  |  |  |
| Female | .9705022 | .0268806 | -1.08 | 0.280 | .9192192 | 1.024646 |

Average values of the dependent variable for all clusters across India(clusters are defined in the NFHS 5 sampling design^15^), which represent average risk scores of the clusters, are plotted as a choropleth graph in Fig. 1 to present a spatial distribution of the high-risk zones. The map was plotted using the software QGIS version 3.40.9.

**Figure 1.**
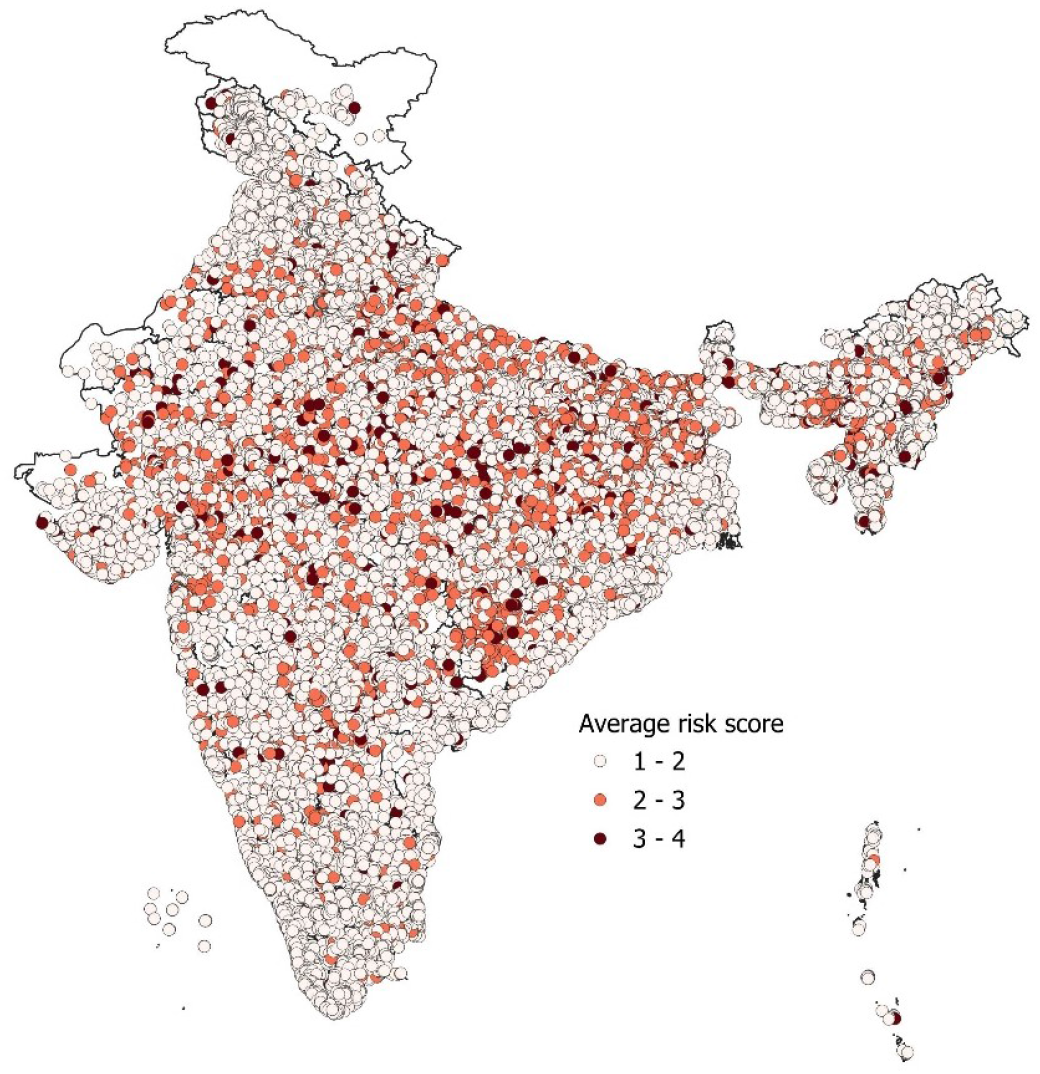
Map showing average risk scores across clusters defined in NFHS-5.

## 3. Results

Bivariate analysis revealed that all examined factors were significantly associated with the outcome variable. However, these associations were unadjusted and do not account for the influence of other variables. Therefore, to better understand the independent effect of each factor, a multivariable ordinal logistic regression was fitted. The results of this adjusted analysis provided the following insights into the associations of maternal and environmental factors with child health outcomes.

### Maternal factors

Maternal age was found to be a significant factor in child health outcomes. Mothers aged 25 to 35 years had a 13% higher risk of adverse outcomes compared to those aged 14 to 24 years, and this risk increased to 27% among mothers aged 35 to 49 years. Birth order also showed significance, with the second and third children having a 15% lower risk of adverse outcomes compared to first-born children. ANC played an important role as well; women who received full ANC had a 14% lower risk of poor outcomes compared to those who did not. Additionally, maternal education had a strong protective effect. Children of mothers with primary education had a 23% lower risk of mortality compared to those whose mothers had no education. This risk reduction increased to 36% for mothers with secondary education and to 52% for those with higher education.

### Environmental factors

Several environmental and socioeconomic factors were found to significantly influence child health outcomes. Children living in rural areas faced a 27% higher risk of adverse outcomes compared to those in urban settings. Use of unsafe cooking fuels was associated with a 17% higher risk of poor outcomes. Institutional delivery was shown to be protective, reducing the risk by 17% compared to non-institutional births. Household wealth was strongly associated with better outcomes.Children from poorer households had a 36% lower risk of adverse outcomes compared to those from the poorest households, and this risk was reduced by as much as 85% for children belonging to the richest quintile. Gender of the child was not found to be significantly associated with the risk of adverse health outcomes, indicating no evidence of gender-based disparity.

The choropleth map of average risk scores (figure 1) shows higher risks of child mortality and morbidity outcomes in India’s central and northern states revealing significant regional disparities in child health. These areas often exhibit higher rates of malnutrition, limited access to healthcare services, and poor sanitation, contributing to adverse child health outcomes. States like Uttar Pradesh, Bihar, Madhya Pradesh, Rajasthan, and Jharkhand have higher concentration of high-risk clusters. These states are part of the Empowered Action Group (EAG) states that were identified by the government of India for special attention due to their socio-economic and demographic challenges which included addressing issues of maternal and child mortality. In contrast, states in southern and western India exhibit lower risk scores.

## 4. Discussion

The ordinal dependent variable used in this analysis provides a simple yet effective means of integrating information on under-five mortality and malnutrition to generate a comprehensive risk score for assessing child health status in a region. It enables the classification of child health status in a region along a continuum, from best (healthy) to worst (mortality), thereby offering deeper insights than traditional metrics such as under-five mortality, infant mortality, or malnutrition alone. For example, from the choropleth graph in Figure 1 various clusters within Rajasthan can be seen to have very high-risk scores. However, as per the NFHS 5 report, the state performs better than 12 States/UTs in terms of under-5 mortality.^15^

Two most important determinants of child health outcome in terms of mortality and child growth factors that emerge from this study are – higher education among the maternal factors and wealth index among the environmental factors. The place of residence is significant with households in rural areas more at risk by a significant 27% as compared to urban areas. Both these results highlight the importance of higher education and poverty reduction especially in rural areas of the country. The gross enrollment rate (GER) in primary school in India is 100% ^18^indicating that almost all children of the eligible age group are enrolled in primary education. However, there is a significant drop to 57.6% at the higher secondary level.^18^ A large portion of students do not transition to or complete higher secondary education.

The global multidimensional poverty index (MPI) is a poverty measure that reflects the multiple deprivations that poor people face in areas of education, health, and living standards. As per the global multidimensional poverty report 2024, India is home to the largest population living in extreme poverty, totaling 234 million people.^19^A twin focus on education and poverty reduction can significantly improve child health outcomes by addressing the root causes of poor health. There is a need for stronger policies and interventions to improve retention, affordability, and accessibility of education at higher levels particularly in rural areas. Education empowers families with knowledge about nutrition, hygiene, and healthcare, thereby enabling them to make informed decisions for their children’s well-being. At the same time, creating jobs and reducing poverty provide families with the financial resources needed to access essential services like medical care, clean water, and nutritious food. For instance, educated parents are more likely to prioritize immunizations and seek timely medical interventions, reducing child mortality and morbidity. A stable family income ensures that children receive adequate nourishment and a healthy living environment, which is essential for their growth and well being.

The risk of mortality is not significantly different between male and female children highlighting gender parity in child health outcomes, a positive indicator for the country. This reflects progress in reducing historical gender-based disparities, particularly in healthcare and nutrition, which have often disadvantaged girls. Such parity may be due to shifting societal norms and effective policy interventions by the government that has been promoting equal healthcare access, nutritional interventions, maternal and child welfare measures, regardless of their gender.

## 5. Conclusion

India has made significant strides in improving child survival rates over the years. However, the country still registered unacceptably high child mortality rates as evident from the data of the latest NFHS 5. Among the G20 nations, a consortium of 19 countries and the European Union that together represent around 85% of the global GDP, over 75% of the global trade, and about two-thirds of the world population (https://www.g20.org),India has the highest neonatal, infant, and under five mortality rates at 20.3, 27 and 32.6 per thousand live births respectively (https://data.worldbank.org). Given the diversity and the complex interaction of many factors there is no one size fit all solution to this problem. One needs to continuously examine data to draw relevant inferences and for timely interventions. Combining child mortality and morbidity to construct the child health outcome as proposed in this study allows better indication of a child’s health status with added sensitivity through progressive health status levels, ranging from best scenario (no malnutrition) to worst scenario (death). Consequently, it enables a more comprehensive assessment of maternal and socioeconomic determinants of child health. In addition, using such combined outcome ensures complete utilization of information available in a survey dataset, irrespective of the mortality status of the child. Identifying the determinants using this approach will not only identify factors that affect child mortality but also lead to a better understanding of the determinants of an improved quality of life with regard to the health of surviving children.

### Limitations

This study proposes an alternative approach to defining and analyzing the health status of children. In estimating the average risk score for a cluster, the ordinal categories of the proposed outcome variable were treated with equal weights. Future research may explore the assignment of differential weights to these categories to enable a more accurate and context-sensitive representation of regional risk scores.

## Data Availability

Data is available in the public domain and can be accessed through the DHS website at https://dhsprogram.com/data.

## Statements and Declarations

### Ethical approval and informed consent statements

No ethical approval was required as this paper is based on secondary analysis of anonymised survey data that is available in the public domain and can be accessed through the DHS website at https://dhsprogram.com/data.

